# Relationship Between Carotid Body Size, Left Atrial Strain, and Non-Invasive Aortic Waveforms in Hypertensive Patients

**DOI:** 10.1101/2025.03.03.25323284

**Authors:** Selen Eşki, Hatice Taşkan, Pelin Meşe Coşkun, Sanan Allahverdiyev, Şeymagül Karaca, Furkan Kırık, Sude Cesaretli, Dilara Akdağ, Salim Yaşar, Erkan Yildirim, Uygar Çağdaş Yüksel, Murat Çelik

**Author notes:** **<u>*Correspondence:*</u>** Selen EŞKİ, M.D., Resident Fellow of Cardiology, Cardiology Clinic, Dr. Yaşar Eryılmaz Doğubeyazıt State Hospital, Ağrı/TÜRKİYE. **Disclosure:** Nothing to disclose.

## Abstract

**Aim:** This study aimed to investigate the effect of carotid body size on target organ damage in patients with hypertension and its potential association with relevant parameters.

**Method:** A total of 49 participants diagnosed with hypertension or newly diagnosed hypertension, who were admitted to the outpatient clinic between January 2023 and April 2024, were included in the study. The size of the carotid body was measured using computed tomography. The study population was categorized into two groups based on a carotid body size threshold of 2.5 mm. Transthoracic echocardiography was performed, and blood pressure measurements and pulse wave analyses were examined non-invasively.

**Results:** The mean age of the participants was 54.93 ± 10.85 years, and 36.7% (n=18) were male. In the group with a larger carotid body size, the left atrial diameter, total vascular resistance, and pulse-adjusted augmentation index (AIx@75) were higher, while the left atrial strain values and cardiac index were lower compared to those in the group with a smaller carotid body size (p <0.05). Multivariate linear regression analysis and ROC analysis were performed to determine the parameters that were significantly associated with carotid body size, and it was revealed that left atrial diameter (AUC: 0.794 (0.667-0.920), p<0.001) and total vascular resistance (AUC: 0.819 (0.700-0.938), p<0.001) were independently associated with carotid body size.

**Conclusion:** This study suggests that processes leading to carotid body enlargement or those caused by this enlargement in patients diagnosed with hypertension, may affect parameters such as left atrial diameter, left atrial strain, AIx, and total vascular resistance, which could serve as indicators of target organ damage.

## Introduction

Hypertension is a chronic and prevalent health condition that is one of the leading risk factors for cardiovascular disease (CVD) and all-cause mortality [1]. Persistently elevated blood pressure negatively impacts individuals’ quality of life and leads to premature death. According to the 2023 report published by the World Health Organization, approximately one-third of the adult population worldwide suffers from high blood pressure [2]. This finding clearly highlights that hypertension is a major global public health issue.

The prevalence of hypertension increases in the presence of risk factors such as advancing age, obesity, sedentary lifestyles, and high salt consumption [3]. Additionally, the often-asymptomatic onset of hypertension makes it challenging to diagnose the disease early and begin effective treatment [4]. Persistently high blood pressure increases the risk of end-stage renal failure and cardiovascular complications, including hemorrhagic or ischemic cerebrovascular accidents, heart failure, coronary artery disease, peripheral artery disease, and sudden death [5]. Elevated blood pressure at younger ages is also associated with cognitive decline, increasing the risk of developing dementia [6].

For all these reasons, understanding the biological mechanisms associated with hypertension is crucial for the development of effective treatment strategies. One of the mechanisms involved in the pathogenesis of hypertension is carotid body hyperactivity [7-10].

The carotid body was first described in the mid-18th century by Albrecht von Haller [11]. This structure is supplied by branches from both the external and internal carotid arteries and is one of the most highly perfused organs in mammals, receiving approximately 2000 mL/min per gram of tissue. The average weight of the carotid body in adults ranges from 18 to 20 milligrams, with its size varying between approximately 2 and 3 millimeters [12]. As a peripheral chemoreceptor, it plays a critical role in hemodynamic regulation in response to arterial oxygen, carbon dioxide, and pH levels [11]. Moreover, carotid body hyperactivity and hypertrophy have been observed in chronic conditions such as hypertension, heart failure, diabetes, and sleep apnea [8,13].

This study aims to investigate the effects of hypertension on the size of the carotid body and explore its potential role in hemodynamic regulation.

## Material and Methods

This retrospective cohort study was conducted on individuals diagnosed with hypertension who visited the cardiology outpatient clinics. Data were collected retrospectively from hospital records between January 2023 and April 2024.

Participants were selected based on specific inclusion and exclusion criteria. The inclusion criteria were defined as patients over 18 years of age who had been diagnosed with systemic hypertension and had undergone CT angiography of the carotid arteries within the past six months. A total of 128 patients who met these criteria were included in the study. The exclusion criteria were as follows: a history of stroke (CVA) or transient ischemic attack (TIA), surgical or percutaneous intervention on the carotid arteries, coronary artery disease, heart failure, atrial fibrillation, moderate to severe valvular diseases detected on echocardiography, chronic obstructive pulmonary disease (COPD), obstructive sleep apnea syndrome (OSAS), or inadequate imaging quality. After exclusions, 49 patients were included in the final analysis.

Carotid artery CT angiography was performed using a 320-slice CT scanner (Aquilion One, Toshiba Medical System, Japan). The dimensions of the carotid body were analyzed using Horos™ v3.3.6 software (www.horosproject.org). The inferomedial area of the carotid bifurcation, where the carotid body is typically located, was examined. The oval structure visible in the arterial phase axial section at this location was identified as the carotid body. To ensure consistency, all participants were analyzed based on the right carotid body. Dimensions less than 2.49 mm were considered normal, while those ≥2.5 mm were classified as enlarged, according to the literature [8,14].

Transthoracic echocardiography recordings of participants were taken using a Philips Epiq 7 device (Philips Medical Systems, Bothell, WA) with a 3.5 MHz transducer. At least three consecutive cycles during the post-expiratory phase were captured. Measurements were analyzed offline by two operators using QLAB 10.1 software (Philips Medical Systems, Bothell, WA, USA). PALS (peak atrial longitudinal strain) and PACS (peak atrial contractile strain), which reflect the reservoir and atrial pump functions, were calculated from strain curves obtained by two-dimensional speckle-tracking echocardiography (2DSTE). Conduit strain was calculated by subtracting PACS from PALS, using the onset of the QRS complex as the reference point.

Blood pressure and pulse wave analyses were measured using a Mobil-O-Graph® device with the ARCSolver algorithm (IEM, Germany; Austrian Institute of Technology, Vienna, Austria). The augmentation index (AIx) was calculated as the ratio of augmented pressure to pulse pressure. Pulse wave velocity (PWV) was calculated based on a mathematical model that considers various parameters. To minimize variability due to heart rate differences, AIx was normalized to a standard heart rate of 75 beats per minute (AIx@75) using the formula: AIx@75 = AIx − 0.39 × (75 − heart rate). Arterial stiffness was evaluated using PWV and AIx@75. Hemodynamic parameters such as stroke volume (SV), cardiac output (CO), and total vascular resistance (TVR) were also assessed.

### Statistical Analysis

Data were analyzed using SPSS (Statistical Package for the Social Sciences) version 25. The normality of the data distributions was tested using the Shapiro-Wilk test. For normally distributed data, descriptive statistics were presented as the mean ± standard deviation, while non-normally distributed data were presented as the median and interquartile range (IQR). Categorical data were analyzed using the chi-square test. To compare independent groups, the Independent Samples T-test was used for normally distributed data, and the Mann-Whitney U test for non-normally distributed data. One-way ANOVA was used to analyze differences in means across groups. Pearson and Spearman correlation analyses were performed to assess relationships between variables. A multivariate regression analysis was conducted to examine factors influencing carotid body dimensions. A receiver operating characteristic (ROC) analysis with the Youden index was used to determine cutoff values for significant variables. Statistical significance was set at p ≤ 0.05.

### Ethical Considerations

The study protocol was approved by the Local Ethics Committee of our Institute (Date: April 24, 2024, Decision No: 2024-223).

## Results

The age range of the patients in the study group was between 31 and 81 years, with a mean age of 54.93 ± 10.85 years. Among the study participants, 36.7% were male (n=18), and 63.3% were female (n=31). The demographic characteristics are shown in Table 1.

**Table 1.** Demographic characteristics of the study group.

| Characteristic | Value |
| --- | --- |
| Age, years | $54.93 \pm 10.85$ |
| BSA, m<sup>2</sup> | $1.91 \pm 0.19$ |
| Male, n (%) | 18 (36.7) |
| Smoking, n (%) | 25 (51) |
| Hypertension, n (%) | 25 (51) |
| Diabetes Mellitus, n (%) | 7 (14.3) |
| Coronary Artery Disease, n (%) | 7 (14.3) |
| Receiving Antihypertensive Treatment, n(%) | 23 (46.9) |

*BSA: body surface area*

The participants were divided into two groups based on carotid body size: those with a carotid body size <2.5 mm (n=22, Group 1) and those with a size ≥ 2.5 mm (n=27, Group 2). In Group 1, the mean carotid body size was 1.91 ± 0.29 mm, while in Group 2, it was 3.14 ± 0.43 mm. HU values from the carotid body and internal carotid artery were compared to evaluate imaging quality. (Table 2.)

**Table 2.** Comparison of parameters according to the carotid body size.

|  | Group 1 (n=22) | Group 2 (n=27) | p-value |
| --- | --- | --- | --- |
| *Tomographic characteristics* |  |  |  |
| Carotid Body Length, mm | 1.91 ± 0.29 | 3.14 ± 0.43 | **< 0.001** |
| Carotid Body HU | 116.00 ± 63.30 | 107.22 ± 42.77 | 0.860 |
| Internal Carotid Artery HU | 314.27 ± 100.39 | 318.59 ± 70.29 | 0.566 |
| Carotid Body HU / ICA HU | 0.38 ± 0.20 | 0.35 ± 0.16 | 0.563 |
| *Echocardiographic parameters* |  |  |  |
| LA diameter, mm | 32,45 ± 3,43 | 36,62 ± 4,10 | **< 0,001** |
| LA area, cm<sup>2</sup> | 14,35 ± 2,35 | 15,60 ± 2,83 | 0,382* |
| LA volume, mL | 31,80 ± 9,09 | 37,01 ± 9,83 | 0,062 |
| *Left atrial strain parameters* |  |  |  |
| LA Global PALS, % | 30.84 ± 11.40 | 23.88 ± 7.70 | **0.014** |
| LA Global PACS, % | 14.10 ± 4.96 | 11.18 ± 4.23 | **0.031** |
| LA Global CDs, % | 16.73 ± 7.52 | 12.69 ± 5.32 | **0.033** |
| LA Stiffness Index | 0.29 ± 0.25 | 0.39 ± 0.23 | **0.015\*** |
| *Peripheral Blood Pressure* |  |  |  |
| SBP, mmHg | 131,63 ± 19,93 | 136,00 ± 16,75 | 0,408 |
| DBP, mmHg | 81,63 ± 14,83 | 89,74 ± 12,33 | **0,042** |
| MAP, mmHg | 104,54 ± 15,37 | 110,81 ± 13,27 | 0,132 |
| PP, mmHg | 50,00 ± 15,81 | 45,14 ± 14,09 | 0,262 |
| *Central Blood Pressure* |  |  |  |
| SBP, mmHg | 133,25 ± 16,19 | 134,95 ± 18,00 | 0,745 |
| DBP, mmHg | 85,15 ± 13,90 | 90,25 ± 13,25 | 0,221 |
| MAP, mmHg | 48,10 ± 14,93 | 44,70 ± 14,20 | 0,445 |
| PP Amplification | 1,31 ± 0,14 | 1,26 ± 0,10 | 0,169 |
| *Hemodynamic Parameters* |  |  |  |
| Stroke Volume, ml | 68,80 ± 13,69 | 63,31 ± 8,33 | 0,091 |
| Cardiac Output, L/dk | 5,20 ± 0,85 | 4,82 ± 0,79 | 0,112 |
| Cardiac Index, L/min/m<sup>2</sup> | 2,79 ± 0,55 | 2,34 ± 0,54 | **0,013\*** |
| TVR, s*mmHg/mL | 1,22 ± 0,18 | 1,56 ± 0,49 | **< 0.001\*** |
| Heart rate, bpm | 77,18 ± 12,28 | 76,51 ± 10,77 | 0,841 |
| *Arterial stiffness parameters* |  |  |  |
| AIx@75, (%) | 24,36 ± 11,77 | 31,11 ± 7,91 | **0,021** |
| PWV, (m/s) | 8,37 ± 1,67 | 8,22 ± 1,35 | 0,728 |
| Reflection Magnitude, (%) | 65,00 ± 10,05 | 65,29 ± 7,23 | 0,905 |
| Augmentation Pressure, mmHg | 12,30 ± 7,96 | 14,12 ± 7,57 | 0,501* |

\*Parameters that did not conform to a normal distribution were compared using the Mann-Whitney U test.
AIx@75: Augmentation Index normalized to a standard heart rate of 75 beats per minute, DBP: Diastolic Blood Pressure, HU: hounsfield unit, ICA: internal carotid artery, MAP: Mean Arterial Pressure, PP: Pulse Pressure, PWV: Pulse Wave Velocity, SBP: Systolic Blood Pressure, TVR: Total Vascular Resistance

Demographic, clinical, and baseline biochemical parameters showed no statistically significant differences between the groups. The use of antihypertensive medications was also comparable between the groups, and blood pressure control was both adequate and similar. However, the spot urine microalbumin-to-creatinine ratio was noticeably higher in Group 2 compared to Group 1, though this difference did not reach statistical significance (14.36 ± 13.86 vs. 27.11 ± 30.00, p = 0.089).

Comparisons of conventional echocardiographic parameters between the groups showed no significant differences in annular aorta, ascending aorta diameters, interventricular septum diastolic diameter (IVSDd), left ventricular posterior wall diastolic diameter (PWDd), and left ventricular diastolic diameter (LVIDd) (for all, p > 0.005). However, the left atrium (LA) diameter was significantly larger in Group 2 (32.45 ± 3.43 mm vs. 36.62 ± 4.10 mm, p <0.001). Although LA area and volume were greater in Group 2, the differences were not statistically significant. In Group 2, left atrial strain parameters showed a tendency to worsen compared to Group 1. The left atrial stiffness index, an early marker of diastolic dysfunction, was significantly higher in Group 2, as shown in Table 2, and Figure 1A and B demonstrate examples of strain echocardiography from both groups.

**Figure 1.**
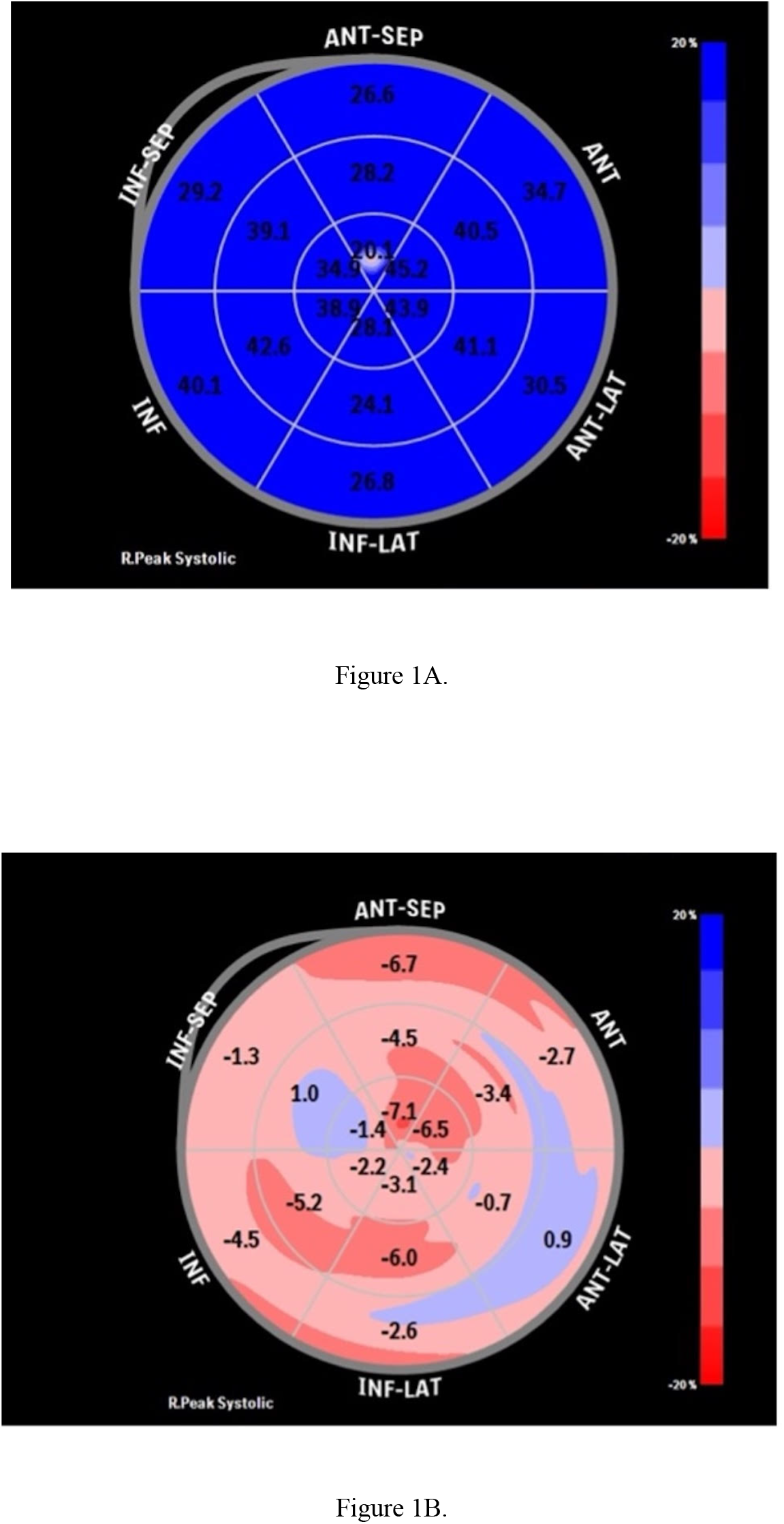
Preserved left atrial strain bull-eye appearance in a participant from Group 1 **(A)**, Impaired left atrial strain bull-eye appearance in a participant from Group 2 **(B)**

Peripheral blood pressure assessments showed no significant differences in systolic blood pressure, mean arterial pressure, or pulse pressure between the groups. However, diastolic blood pressure was significantly different (p=0.042). There were no significant differences in central blood pressures or pulse pressure amplification between the groups. The cardiac index (p=0.006) and total vascular resistance (p=0.004) showed significant differences between the groups. (Table 2) AIx@75 was significantly higher in Group 2 (p=0.021). Although other parameters indicating arterial stiffness, such as PWV, augmentation pressure, and reflection magnitude, tended to worsen in Group 2, no statistically significant differences were observed. (Table 2)

The parameters found to be significant in the univariate correlation analysis are presented in the table 3. Multivariate linear regression analysis revealed that LA diameter, total vascular resistance, cardiac output, and the spot urine microalbumin-to-creatinine ratio were independently associated with carotid body size (Table 4) In the regression analysis, 42.9% of the abnormal increase in carotid body size can be attributed to these variables.

**Table 3.** Univariate correlation analysis.

|  | **r value** | **p value** |
| --- | --- | --- |
| Spot urine microalbumin/creatinine ratio | 0.329 | 0.021 |
| IVSDd, mm | 0.292 | 0.042 |
| LA, mm | 0.451 | 0.001 |
| Mitral A velocity, m/s | 0.326 | 0.022 |
| Mitral e' velocity, cm/s | -0.283 | 0.048 |
| Pulse pressure, mmHg | -0.293 | 0.041 |
| Stroke volume, ml | -0.282 | 0.05 |
| Cardiac output, L/min | -0.296 | 0.039 |
| Cardiac index, L/min/m<sup>2</sup> | -0.401 | 0.004 |
| TVR, s*mmHg/ml | 0.377 | 0.008 |
| AIx75, % | 0.297 | 0.038 |

**Table 4.** Multivariate linear regression analysis.

|  | **B (%95 CI)** | **t** | **p value** | **Zero order** | **Partial order** |
| --- | --- | --- | --- | --- | --- |
| LA diameter, mm | 0.640 (0.025-0.103) | 3.342 | 0.002 | 0.442 | 0.472 |
| TVR,s*mmHg/ml | 0.553 (0.161-0.944) | 2.856 | 0.007 | 0.348 | 0.416 |
| Spot Urine Microalbumin-to-Creatinine Ratio | 0.009 (0.002-0.015) | 2.611 | 0.013 | 0.379 | 0.386 |
| Cardiac Output, L/min | -0.240 ((-0.36) -(-0.281)) | 2.381 | 0.022 | -0.281 | -0.356 |

*Adj.R<sup>2</sup>:0.429 SE:0.546 F:5.668 p:0.022*

In the ROC curve analysis performed to determine the accuracy, performance, and cut- off values of the variables found to be significant in the regression analysis, LA diameter and TVR were found to be statistically significant. However, cardiac output (p=0.87) and the spot urine microalbumin-to-creatinine ratio (p=0.77) did not show statistically significant differences. The cut-off values for LA diameter and TVR were determined using the Youden index in the ROC analysis. The results showed that when the LA diameter is ≥ 34.50 mm, a carotid body size of 2.5 mm or more is observed with 70.3% sensitivity and 81.8% specificity (AUC: 0.794 (0.667-0.920), p<0.001). Similarly, when TVR is ≥ 1.35 s*mmHg/ml, a carotid body size of 2.5 mm or more is associated with 66.6% sensitivity and 81.8% specificity (AUC: 0.819 (0.700-0.938), p<0.001). (Figure 2A and B)

**Figure 2.**
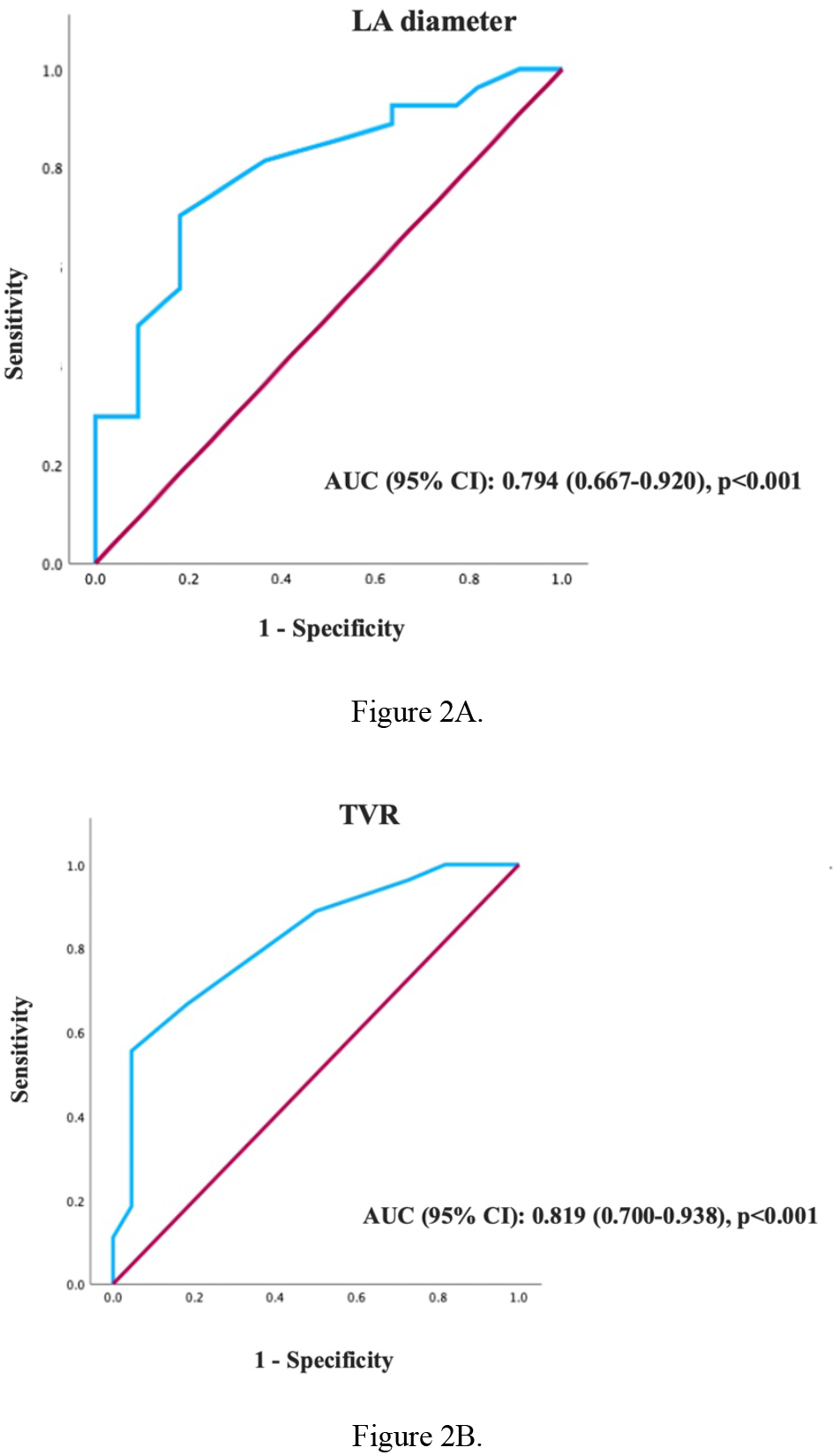
ROC curve for estimating carotid body size ≥ 2.5mm based on measured LA Diameter (**A**) and TVR (**B**)

## Discussion

To the best of our knowledge, no study has investigated the relationship between carotid body size and LA strain and non-invasive arterial stiffness parameters in the assessment of target organ damage. We found that patients diagnosed with hypertension with a carotid body size of 2.5 mm or greater had more impaired left atrial strain and arterial stiffness parameters (mainly TVR and AIx@75), which could serve as indicators of target organ damage, compared to those without.

The study by R.P. Nguyen et al [14] was the first to evaluate the carotid body using CT angiography. They found that the average size of the right carotid body was 2.4 × 2.0 (± 0.8 × 0.6) mm, and the left carotid body was 2.2 × 2.1 (± 0.7 × 0.5) mm [14]. Constant stimulation can make the carotid body more sensitive and cause it to enlarge. An enlarged carotid body is linked to increased sympathetic activity and chemoreflex responses, and it has been observed in conditions such as COPD, OSA, hypertension, heart failure, and diabetes [13]. In a study by J.A. Cramer et al [8], carotid body sizes were compared with different diseases. They found that the average size in healthy individuals was 2.3 mm, while patients with hypertension, heart failure, and diabetes had 20-25% larger carotid bodies [8]. Based on the literature, we set the threshold value for the right carotid body size at 2.5 mm using CT angiography in our study group.

Target organ damage is generally observed in patients with prolonged or severe hypertension, but it may also occur at lower blood pressure levels. However, it remains unclear which patients develop damage earlier or more severely. In our study, we investigated the relationship between hypertension and carotid body size and found that enlargement of the carotid body may be associated with early indicators of cardiovascular target organ damage.

Although PWV is considered the gold standard for evaluating central arterial stiffness, AIx has been shown to reflect systemic arterial stiffness [15]. While it is not yet widely used in clinical practice, many studies have demonstrated that AIx could be an independent predictor of cardiovascular events [16, 17]. In the Conduit Artery Function Evaluation (CAFE) study, central pulse pressure and AIx were found to be independent predictors of all-cause mortality in hypertensive patients [18]. In a study by Saba et al [19], AIx values were associated with left ventricular hypertrophy in normotensive patients. In patients with resistant hypertension, increased arterial stiffness, impaired endothelial function, and elevated AIx values have been considered indicators of cardiovascular events [20]. In our study, contrary to expectations, no difference was observed between the two groups regarding PWV, which we attribute to the adequate control of blood pressure in most participants during follow-up. However, patients with an increased carotid body size showed higher AIx@75 values and TVR. These findings suggest that AIx reflects not only central but also systemic vascular stiffness and may be associated with microvascular damage, aligning with the literature.

Aortic stiffness and the onset of hypertension remain subjects of debate in the academic world. Elevated blood pressure is thought to stress the aortic wall, leading to elastin loss and increased collagen, which results in aortic stiffness [21]. However, some studies suggest that increased aortic stiffness may predict future hypertension [22]. It is known that increased aortic stiffness can lead to a reduction in global longitudinal strain in the left ventricle over time. However, early diastolic filling may be affected before this deterioration becomes apparent [23]. Consistent with the literature, our study found that hypertensive patients without systolic dysfunction, but with a larger carotid body size, exhibited diastolic dysfunction and higher AIx@75 values.

Hypertension tends to affect diastolic function before systolic function, and impaired left ventricular diastolic function can increase LA load through elevated filling pressures [24]. Functional changes in the LA occur before structural changes, leading to reduced reservoir function [25]. Over time, structural alterations such as atrial dilation, increased volume load, and left ventricular hypertrophy develop. Strain and strain rate, measured using 2DSTE, are more sensitive in detecting early functional changes in the LA compared to conventional echocardiographic measurements [26]. In our study, hypertensive patients with larger carotid body sizes exhibited lower PALS, PACS, and conduit strain values. This suggests that LA dysfunction may be associated with carotid body enlargement in hypertensive patients. It is known that LA strain deteriorates over time, and LA diameter and volume increase in hypertensive patients [27, 28]. To our knowledge, there are no studies in the literature that explore the relationship between carotid body size and LA strain. The study by Yixiao Zhao et al [29] in 2020 showed that the LA stiffness index is independently associated with urinary microalbumin excretion, PWV, and left ventricular hypertrophy in hypertensive patients. This suggests that the LA stiffness index could be associated with systemic vascular dysfunction and endothelial damage, helping in the early detection of organ damage. In our study, we excluded patients with heart failure, and all participants had preserved left ventricular systolic function. However, patients with larger carotid bodies exhibited more significant left ventricular diastolic dysfunction and higher LA stiffness indices.

Microalbuminuria is considered an early indicator of hypertensive kidney damage and is known to be associated with endothelial dysfunction [30-33]. In our study, patients with larger carotid body sizes had higher morning first-void urine microalbumin levels compared to the other group, and the spot urine microalbumin-to-creatinine ratio was also higher. Since our sample size was not large enough, we believe this difference did not reach statistical significance. However, both univariate and multivariate analyses showed a positive correlation between carotid body size and the microalbumin-to-creatinine ratio. This finding may be an early sign of subclinical organ damage in individuals with a diagnosis of hypertension and a carotid body size of 2.5 mm or greater. However, it is obvious that large-scale, prospective studies are needed to confirm this.

In patients whose blood pressure cannot be controlled with conventional treatments, invasive therapies may be considered. An increase in carotid body size is known to be associated with elevated sympathetic activity and hypertension. Studies in the literature suggest that carotid body excision may be an effective method for managing resistant hypertension [34-37]. Additionally, resection of the superior cervical ganglion or medical interventions using alpha- 1 adrenergic receptor antagonists have been shown to significantly reduce increased chemoreflex responses [38]. Future studies may show that surgical and medical treatments could become more widely used in patients with carotid body hypertrophy and increased chemoreflex responses.

### Limitations

Our study has several limitations. Firstly, since this was a single-center retrospective study, its results should be considered within the context of the typical limitations associated with retrospective study. Secondly, due to the relatively small sample size, which may have affected the statistical power of our analyses, the results of this study should be validated in larger patient cohorts and through multicenter studies.

## Data Availability

The data that support the findings of this study are available from the corresponding author upon reasonable request.

## Conclusion

In our study, the group with larger carotid body sizes showed impaired LA strain, diastolic dysfunction, increased AIx and TVR, and higher levels of spot urine microalbumin compared to the group with smaller carotid bodies. This was observed even in the absence of resistant hypertension or consistently high blood pressure. The findings of this study suggest that carotid body enlargement could be a useful clinical marker for the early detection of target organ damage and contribute to the development of more tailored treatment strategies in individuals with hypertension. Future prospective studies involving a broader range of populations could improve the generalizability of these findings. Additionally, more in-depth molecular investigations into the carotid body’s pathophysiological role could offer valuable insights into its involvement in hypertension management.

## References

1. Mills, K.T., A. Stefanescu, and J. He, The global epidemiology of hypertension. Nature Reviews Nephrology, 2020. 16(4): p. 223–237.

2. Organization, W.H., Global report on hypertension: the race against a silent killer. 2023.

3. Chow, C.K., et al., Prevalence, awareness, treatment, and control of hypertension in rural and urban communities in high-, middle-, and low-income countries. Jama, 2013. 310(9): p. 959–968.

4. Viera, A.J., Screening for hypertension and lowering blood pressure for prevention of cardiovascular disease events. Medical Clinics, 2017. 101(4): p. 701–712.

5. Rapsomaniki, E., et al., Blood pressure and incidence of twelve cardiovascular diseases: lifetime risks, healthy life-years lost, and age-specific associations in 1· 25 million people. The Lancet, 2014. 383(9932): p. 1899–1911.

6. Daugherty, A.M. Hypertension-related risk for dementia: A summary review with future directions. in Seminars in Cell & Developmental Biology. 2021. Elsevier.

7. Nair, S., et al., CT angiography in the detection of carotid body enlargement in patients with hypertension and heart failure. Neuroradiology, 2013. 55(11): p. 1319–22.

8. Cramer, J., et al., Carotid body size on CTA: correlation with comorbidities. Clinical radiology, 2014. 69(1): p. e33–e36.

9. Felippe, I.S., et al., Commonalities and differences in carotid body dysfunction in hypertension and heart failure. The Journal of Physiology, 2023. 601(24): p. 5527–5551.

10. Shell, B., K. Faulk, and J.T. Cunningham, Neural control of blood pressure in chronic intermittent hypoxia. Current hypertension reports, 2016. 18: p. 1–9.

11. Kumar, P. and N.R. Prabhakar, Peripheral chemoreceptors: function and plasticity of the carotid body. Compr Physiol, 2012. 2(1): p. 141–219.

12. Iturriaga, R., M.P. Oyarce, and A.C.R. Dias, Role of Carotid Body in Intermittent Hypoxia-Related Hypertension. Curr Hypertens Rep, 2017. 19(5): p. 38.

13. Thakkar, P., et al., Carotid body: an emerging target for cardiometabolic co-morbidities. Experimental Physiology, 2023. 108(5): p. 661–671.

14. Nguyen, R., et al., Carotid body detection on CT angiography. American Journal of Neuroradiology, 2011. 32(6): p. 1096–1099.

15. Seravalle, G. and G. Grassi, Carotid baroreceptor stimulation in resistant hypertension and heart failure. High Blood Pressure & Cardiovascular Prevention, 2015. 22: p. 233–239.

16. Shimizu, M. and K. Kario, Role of the augmentation index in hypertension. Therapeutic advances in cardiovascular disease, 2008. 2(1): p. 25–35.

17. Chirinos, J.A., et al., Aortic pressure augmentation predicts adverse cardiovascular events in patients with established coronary artery disease. Hypertension, 2005. 45(5): p. 980–985.

18. Investigators, C., et al., Differential impact of blood pressure–lowering drugs on central aortic pressure and clinical outcomes: principal results of the Conduit Artery Function Evaluation (CAFE) study. Circulation, 2006. 113(9): p. 1213–1225.

19. Saba, P.S., et al., Relation of arterial pressure waveform to left ventricular and carotid anatomy in normotensive subjects. Journal of the American College of Cardiology, 1993. 22(7): p. 1873–1880.

20. Alsharari, R., G.Y. Lip, and A. Shantsila, Assessment of arterial stiffness in patients with resistant hypertension: additional insights into the pathophysiology of this condition? American Journal of Hypertension, 2020. 33(2): p. 107–115.

21. Mitchell, G.F., Arterial stiffness and hypertension: chicken or egg? Hypertension, 2014. 64(2): p. 210–214.

22. Kaess, B.M., et al., Aortic stiffness, blood pressure progression, and incident hypertension. Jama, 2012. 308(9): p. 875–881.

23. Boutouyrie, P., et al., Arterial stiffness and cardiovascular risk in hypertension. Circulation research, 2021. 128(7): p. 864–886.

24. Nadruz, W., A.M. Shah, and S.D. Solomon, Diastolic dysfunction and hypertension. Medical Clinics, 2017. 101(1): p. 7–17.

25. Hoit, B.D., Left atrial size and function: role in prognosis. Journal of the American College of Cardiology, 2014. 63(6): p. 493–505.

26. Braunauer, K., et al., Early detection of cardiac alterations by left atrial strain in patients with risk for cardiac abnormalities with preserved left ventricular systolic and diastolic function. The international journal of cardiovascular imaging, 2018. 34:. 701–711.

27. Ikejder, Y., et al., Impact of arterial hypertension on left atrial size and function. BioMed Research International, 2020. 2020(1): p. 2587530.

28. Miljković, T., et al., Left atrial strain as a predictor of left ventricular diastolic dysfunction in patients with arterial hypertension. Medicina, 2022. 58(2): p. 156.

29. Zhao, Y., et al., Left atrial stiffness index as a marker of early target organ damage in hypertension. Hypertension Research, 2021. 44(3): p. 299–309.

30. Cohuet, G. and H. Struijker-Boudier, Mechanisms of target organ damage caused by hypertension: therapeutic potential. Pharmacology & therapeutics, 2006. 111(1): p. 81–98.

31. Klag, M.J., et al., Blood pressure and end-stage renal disease in men. New England Journal of Medicine, 1996. 334(1): p. 13–18.

32. Pedrinelli, R., et al., Microalbuminuria and endothelial dysfunction in essential hypertension. The Lancet, 1994. 344(8914): p. 14–18.

33. Shantouf, R.S., et al., Page. Should Microalbuminuria Ever Be Considered as a Renal Endpoint in Any Clinical Trial? American Journal of Nephrology, 2010. 31(5): p. 457–457.

34. Winter, B. and B.J. Whipp, Immediate effects of bilateral carotid body resection on total respiratory resistance and compliance in humans, in Post-genomic perspectives in modeling and control of breathing. 2004, Springer. p. 15–21.

35. Narkiewicz, K., et al., Unilateral carotid body resection in resistant hypertension: a safety and feasibility trial. JACC: Basic to Translational Science, 2016. 1(5): p. 313–324.

36. NAKAYAMA, K., Surgical removal of the carotid body for bronchial asthma. Diseases of the Chest, 1961. 40(6): p. 595–604.

37. Fudim, M., et al., Effects of carotid body tumor resection on the blood pressure of essential hypertensive patients. Journal of the American Society of Hypertension, 2015. 9(6): p. 435–442.

38. Felippe, I.S.A., et al., The sympathetic nervous system exacerbates carotid body sensitivity in hypertension. Cardiovasc Res, 2023. 119(1): p. 316–331.

